# Patient experiences of chronic postsurgical pain after caesarean delivery: an inductive thematic analysis

**DOI:** 10.1101/2025.09.15.25335765

**Authors:** S Ciechanowicz, P Callihan, G Michel, DM Panelli, B Carvalho, P Sultan

**Affiliations:** Department of Anesthesiology, Perioperative and Pain Medicine, Stanford University School of Medicine, Stanford, CA, USA; Department of Anaesthesia and Perioperative Medicine, University College London Hospitals NHS Foundation Trust, UK; Department of Surgery and Cancer, Faculty of Medicine, Imperial College London, London, UK

**Keywords:** Biopsychosocial, Chronic postsurgical pain, Caesarean delivery, SPACE framework, patient experiences

## Abstract

**Background:** Chronic postsurgical pain after caesarean delivery affects 10-20% of women at 3-6 months postpartum, yet its broader impact on recovery is underexplored. This study examined lived experiences of chronic postsurgical pain and identified key domains of impact.

**Methods:** Twenty-four women with self-reported pain at 3-6 months after intrapartum or planned caesarean delivery were recruited from two prospective studies. Semi-structured interviews, conducted in English or Spanish via secure video call, were transcribed and analysed using inductive reflexive thematic analysis.

**Results:** Participants described a multidimensional, interconnected symptom burden. Pain persisted or worsened unpredictably, interfering with mobility, infant care, and daily life. Poor sleep and fatigue compounded distress. Cognitive and affective disruptions, including anxiety and fear, were common. Many avoided strong analgesics due to concerns about alertness or breastfeeding. Participants sometimes reported feeling dismissed or unsupported by healthcare professionals. Ten themes were identified: pain and sensory disruption; functional limitations and fatigue; interference with infant care and identity; psychological distress and cognitive load; sleep disruption; control and coping; intimacy and embodied recovery; healthcare gaps; peer and online normalisation; and reflections on future health.

**Conclusions:** Chronic pain after caesarean rarely occurs in isolation. Inter-related symptoms across sleep, pain, affect, cognition, and energy domains contribute to the lived experience of chronic caesarean delivery pain. These findings align with the multidomain SPACE-Postpartum framework, and support its further evaluation as a model for understanding and predicting postpartum pain outcomes.

**Highlights:**

- Chronic postsurgical pain (CPSP) after caesarean delivery is a multidimensional burden beyond pain intensity.
- Ten CPSP themes spanned physical, emotional, social, and care impacts.
- Symptoms reinforced each other, compounding recovery challenges.
- Narratives align with the SPACE (Sleep, Pain, Affect, Cognition, Energy) - Postpartum framework of domains.

## Introduction

Caesarean delivery (CD) is the most common inpatient surgical procedure worldwide, accounting for approximately one in four births^1^. In a US survey of new mothers, 79% of those who underwent CD experienced incision pain in the first 2 months (one-third rating it a major problem)^2^. Uncontrolled post-caesarean pain can significantly interfere with early maternal recovery and caregiving. Studies have linked severe postoperative pain to delayed ambulation and breastfeeding initiation^3^, and individuals report that higher pain levels hinder infant care and bonding^4^. Beyond the acute period, a subset of patients develop chronic postsurgical pain (CPSP) after CD, defined as pain persisting beyond 3 months. The true incidence of chronic pain after CD is unknown, but studies suggest approximately 10-20% of women report persistent incision pain at 3-6 months postpartum^5^. Around 5-10% continue to have pain at 12 months, although some populations report rates as high as 18% at one year after CD.^6^ A recent meta-analysis estimated that roughly 1 in 10 patients who undergo CD experience persistent pain severe enough to interfere with daily functioning, quality of life, work, self-care, or social activities months after delivery^5^.

This study used qualitative interviews to explore the lived experiences of individuals with CPSP after CD. The primary aim was to identify key symptoms, impacts, and coping strategies through an inductive reflexive thematic analysis, generating a rich, experience-based understanding of postpartum impact of CPSP after CD.

## Methods

Ethical approval was obtained from the University IRB (Protocols #71824 and #68558). Participants were identified from two ongoing prospective studies. Women aged ≥18 years old, who had undergone a planned or unplanned CD and were still experiencing pain related to their CD beyond 3 months postpartum, were eligible for enrolment. Eligibility criteria included participants who had undergone CD within the past 3-6 months, were aged ≥18 years, and were able to participate in an interview in either English or Spanish. Both primiparous and multiparous individuals were included, as were those who received spinal, epidural, or general anaesthesia. Variation was allowed in chronic pain history, psychiatric history, perinatal complications, insurance status, and neonatal outcomes, to ensure a diverse sample. Individuals with current or past substance misuse, ongoing opioid therapy, or pre-existing mental health conditions were not excluded. Participants were eligible regardless of whether the pregnancy was singleton or multiple. Exclusion criteria included inability to provide informed consent, language other than English or Spanish, major postpartum complications that precluded meaningful participation, and neonatal demise or intrauterine fetal death (IUFD). Major complications were defined as severe medical or surgical morbidity requiring prolonged hospitalisation, intensive care unit admission, or resulting in substantial cognitive or physical impairment interfering with interview participation. For study #71824 general anaesthesia and vertical midline incision were also exclusion criteria.

Purposive sampling was used to capture a range of postpartum pain and recovery experiences. Specifically, we aimed to recruit over 20 individuals, to reach conceptual saturation. The study followed COREQ guidelines (Table 1). Semi-structured interviews were conducted via secure video call between October 2023 and January 2025, at 3-6 months postpartum. Interviews explored pain experiences, recovery, and symptom impacts. Trained female researchers conducted the interviews. S.C conducted English and G.M conducted Spanish interviews. Both interviewers had prior experience of qualitative interview methods. Participants were introduced to the interviewers via the study recruitment process, and were briefed on the aims of the study to improve postpartum pain management.

**Table 1.** COREQ Checklist.

| Domain | Item | COREQ Question | Your Study |
| --- | --- | --- | --- |
| Research team and reflexivity | Interviewer/facilitator | Which author conducted the interview or focus group? | SC, GM |
| Research team and reflexivity | Credentials | What were the researcher's credentials? | MD, |
| Research team and reflexivity | Occupation | What was their occupation at the time of the study? | Clinical Fellow |
| Research team and reflexivity | Gender | Was the researcher male or female? | Female |
| Research team and reflexivity | Experience and training | What experience or training did the researcher have? | 15+ years clinical experience; qualitative research training during PhD |
| Study design | Methodological orientation | What methodological orientation was stated to underpin the study? | Reflexive thematic analysis was conducted inductively. After coding, themes were compared to the SPACE framework as an interpretive lens for integration with cohort and population-level findings. |
| Study design | Sampling | How were participants selected? | Purposive sampling of individuals with chronic pain 3–6 months post-caesarean |
| Study design | Method of approach | How were participants approached? | Participants recruited via prior cohort participation and contacted directly |
| Study design | Sample size | How many participants were in the study? | 24 (19 English, 5 Spanish) |
| Study design | Non-participation | How many people refused to participate or dropped out? | None reported |
| Study design | Setting of data collection | Where was the data collected? | Remote interviews via video call |
| Study design | Presence of non-participants | Was anyone else present besides the participants and researchers? | No |
| Study design | Description of sample | What are the important characteristics of the sample? | Post-caesarean participants with chronic pain (3–6 months), mix of parity, language |
| Study design | Interview guide | Were questions, prompts, guides provided by the authors? Was it pilot tested? | Yes; semi-structured guide iteratively refined. Not pilot-tested formally |
| Study design | Repeat interviews | Were repeat interviews carried out? | No |
| Study design | Audio/visual recording | Did the research use audio or visual recording to collect the data? | Yes, all were audio recorded |
| Study design | Field notes | Were field notes made during and/or after the interview or focus group? | No field notes taken |
| Study design | Duration | What was the duration of the interviews or focus group? | 10–30 minutes per interview |
| Study design | Data saturation | Was data saturation discussed? | Yes, saturation noted after ~20 transcripts |
| Study design | Transcripts returned | Were transcripts returned to participants for comment and/or correction? | No |
| Analysis and findings | Number of data coders | How many data coders coded the data? | SC, PC, GM, PS |
| Analysis and findings | Description of the coding tree | Did authors provide a description of the coding tree? | Yes, the coding tree was structured to capture health-related quality of life impacts and mapped onto the SPACE framework domains, with inductively derived subthemes |
| Analysis and findings | Derivation of themes | Were themes identified in advance or derived from the data? | Themes were developed inductively from the data, then compared and aligned with the SPACE framework domains to allow integration. Subthemes emerged inductively within each domain |
| Analysis and findings | Software | What software, if applicable, was used to manage the data? | Data were coded manually and supported by AI-assisted text management tools; final coding outputs were organised in Excel and Word |
| **Analysis and findings** | Participant checking | Did participants provide feedback on the findings? | No |
| **Analysis and findings** | Quotations presented | Were participant quotations presented to illustrate the themes/findings? | Yes, quotes presented for each domain |
| **Analysis and findings** | Data and findings consistent | Was there consistency between the data presented and the findings? | Yes, strong alignment of data and findings |
| **Analysis and findings** | Clarity of major themes | Were major themes clearly presented in the findings? | Yes |
| **Analysis and findings** | Clarity of minor themes | Is there a description of diverse cases or discussion of minor themes? | Yes, minor and divergent experiences described |

A flexible interview guide was developed to explore recovery experiences while allowing participants to raise any concerns (Appendix 1). Prompts included: “Can you describe your experience of pain after caesarean delivery?”; “How has pain affected your daily life since delivery?”. Follow-up probes encouraged elaboration. Interviews were conducted via secure video conferencing (Zoom) and lasted 10-30 minutes, until conceptual saturation was achieved (3 consecutive interviews with no new themes discussed). All were audio-recorded with permission, transcribed verbatim, and where necessary, translated using Grain transcription platform (https://grain.com). Transcripts were checked for accuracy against audio files. Thematic analysis followed Braun and Clarke’s inductive approach^7,8^. In brief, this involves six phases: (1) familiarisation with the data, (2) generating initial codes, (3) searching for themes, (4) reviewing themes, (5) defining and naming themes, and (6) producing the report.

Coding was inductive, by 4 investigators (S.C, P.C, G.M, P.S) iteratively refined across six rounds (Appendix 2), and analysed using NVivo 14 (version 14.24.3, QSR International, Melbourne, Australia). Transcripts were initially double-coded by all four investigators to develop a shared coding framework. Coding consistency was achieved iteratively through six rounds of discussion and refinement, after which the majority of transcripts were coded by one or two investigators with ongoing peer debriefing. Following Braun and Clarke’s six-phase reflexive thematic analysis, we coded inductively, initially with dual coding, iterative comparison, and team reflexivity. Codes were grouped into higher-order domains through constant comparison. Themes were refined via peer debriefing and audit trails, enhancing trustworthiness while recognising coding as an active, reflexive process rather than a mechanical task.

## Results

### Overview of Participants

Twenty-four women participated (19 English-speaking and 5 Spanish-speaking). Participants were aged between 23 and 40, and CD indications included primary and repeat, and scheduled and unplanned/emergency procedures. All participants received neuraxial anaesthesia (spinal or epidural), and none reported anaesthesia complications. At the time of the interview, four had a prior history of chronic pain or opioid use, two had sought medical care for pain issues postpartum, and three experienced wound infections. Clinical and psychosocial diversity provided a robust foundation for exploring pain experiences and impacts.

Key concepts were identified through seven iterative rounds of thematic analysis, with agreement between investigators. Thematic analysis yielded ten major themes, derived from subdomains (Figure 1, Appendix 2). These themes reflect the multidimensional burden of CPSP, spanning physical, psychological, cognitive, social, and healthcare system domains. Relational and healthcare context influences are also explored. Postpartum pain symptoms, impacts, and coping strategies were grouped according to themes:

**Figure 1:**
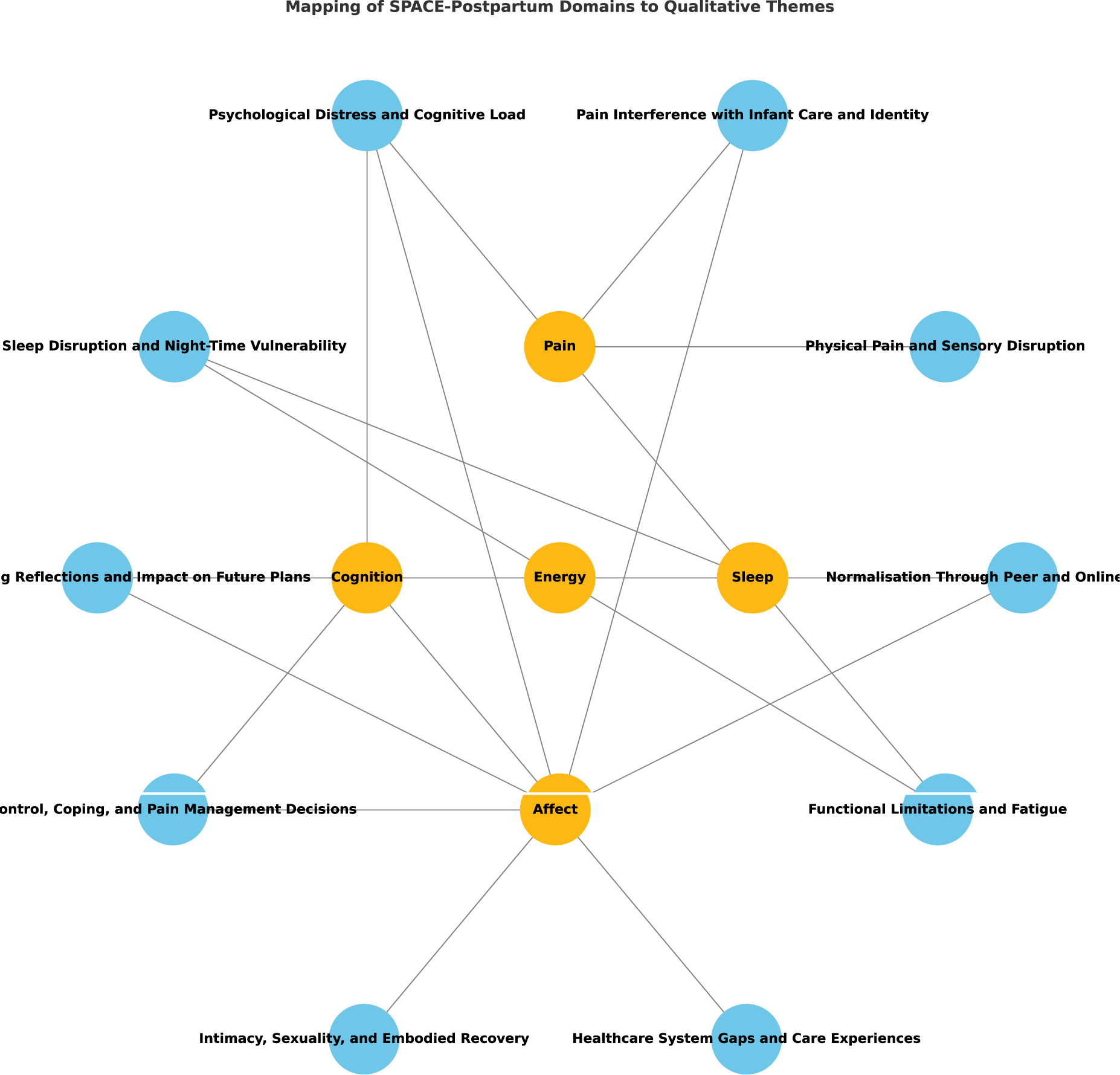
Mapping of SPACE-Postpartum framework to 10 qualitative themes. Mapping was performed after inductive theme generation.

#### 1. Physical Pain and Sensory Disruption

Participants consistently described the acute postoperative period as marked by severe pain. This pain often exceeded expectations and interfered with basic movements such as getting out of bed, walking, or holding the infant.

Descriptions included altered sensations, dull aches, sharp or burning sensations, scar hypersensitivity, and intermittent pain, often aggravated by activity or pressure. The pain recovery is described to initially improve but then plateau. Examples are outlined in Table 2.

**Table 2.**
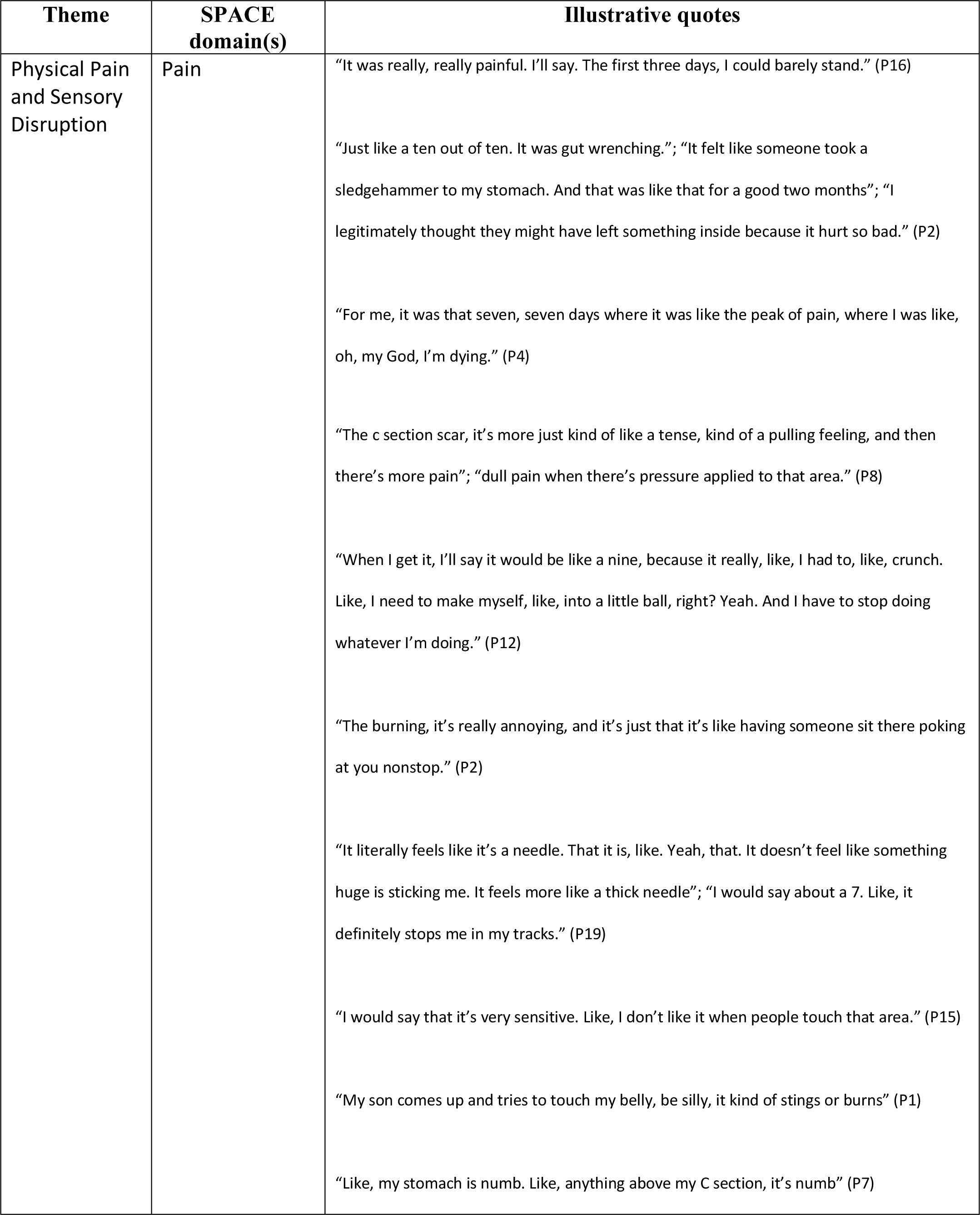
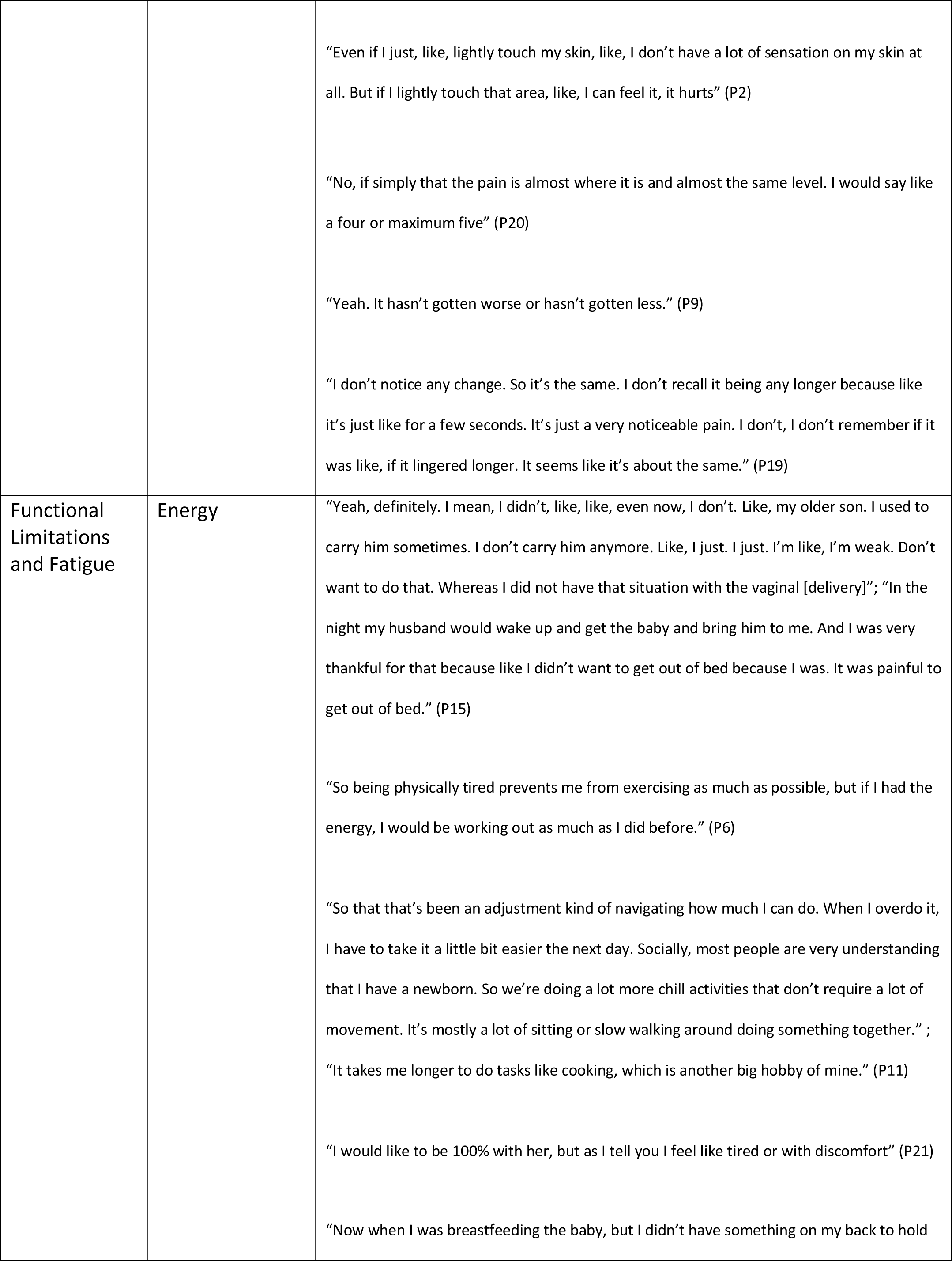
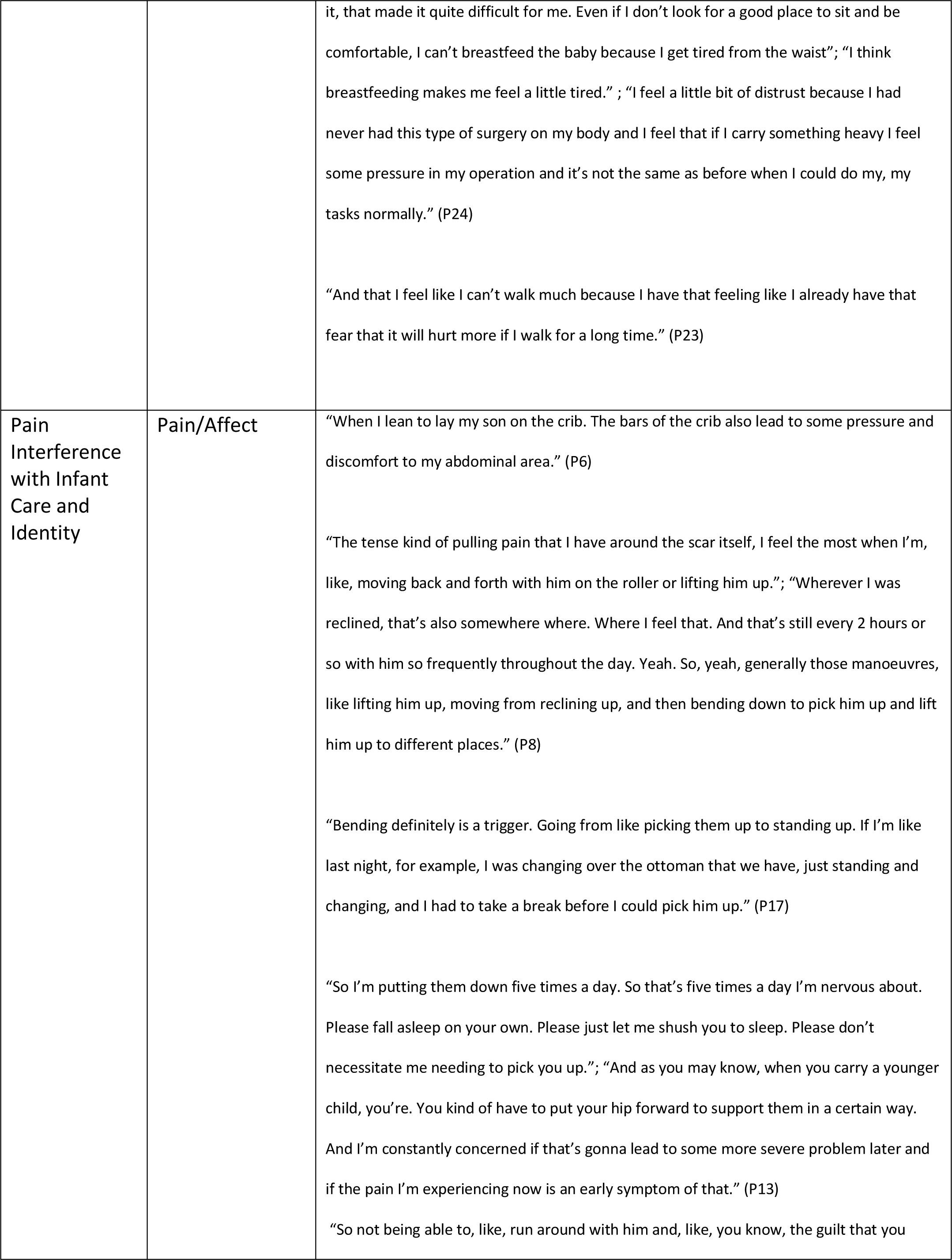
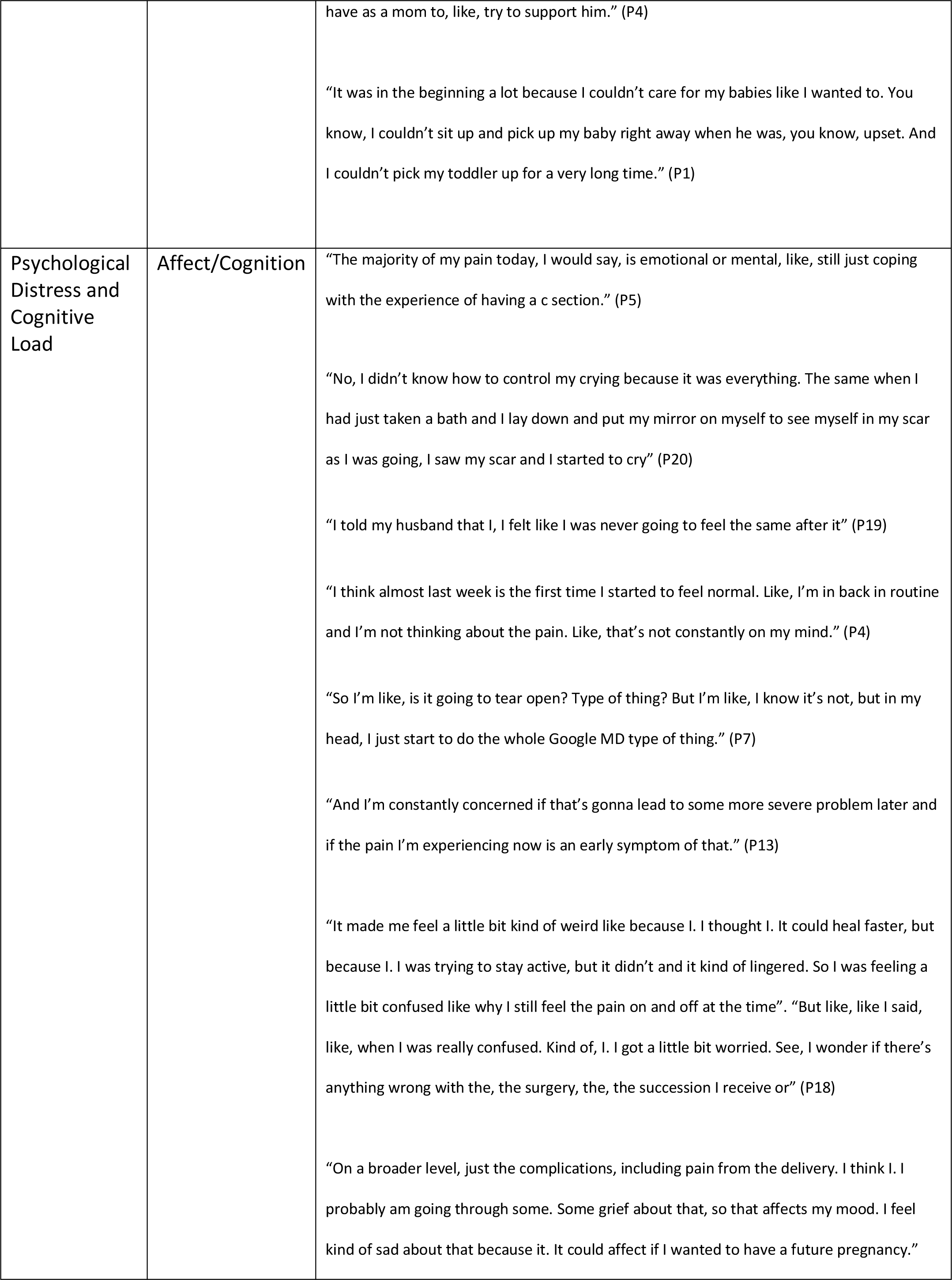
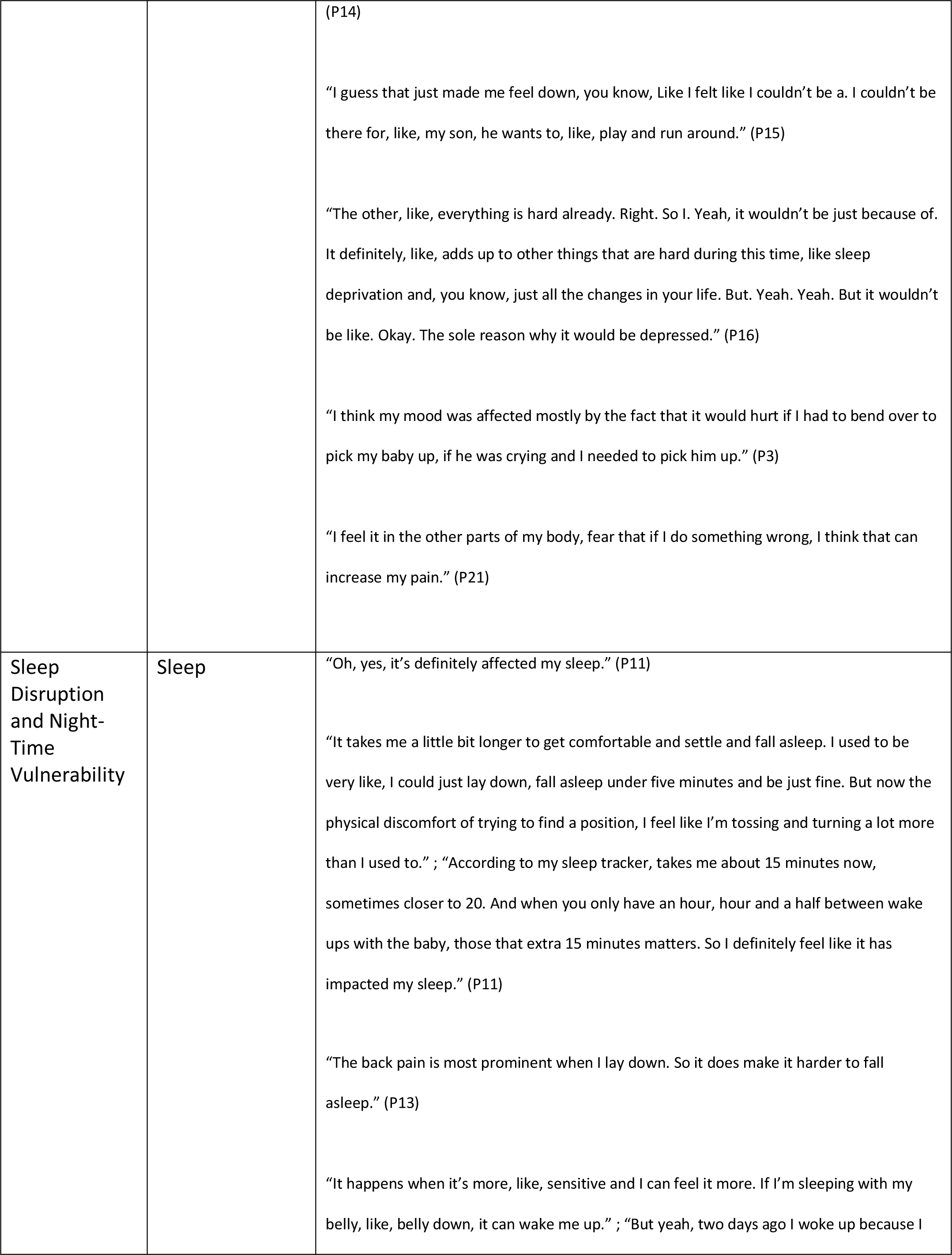
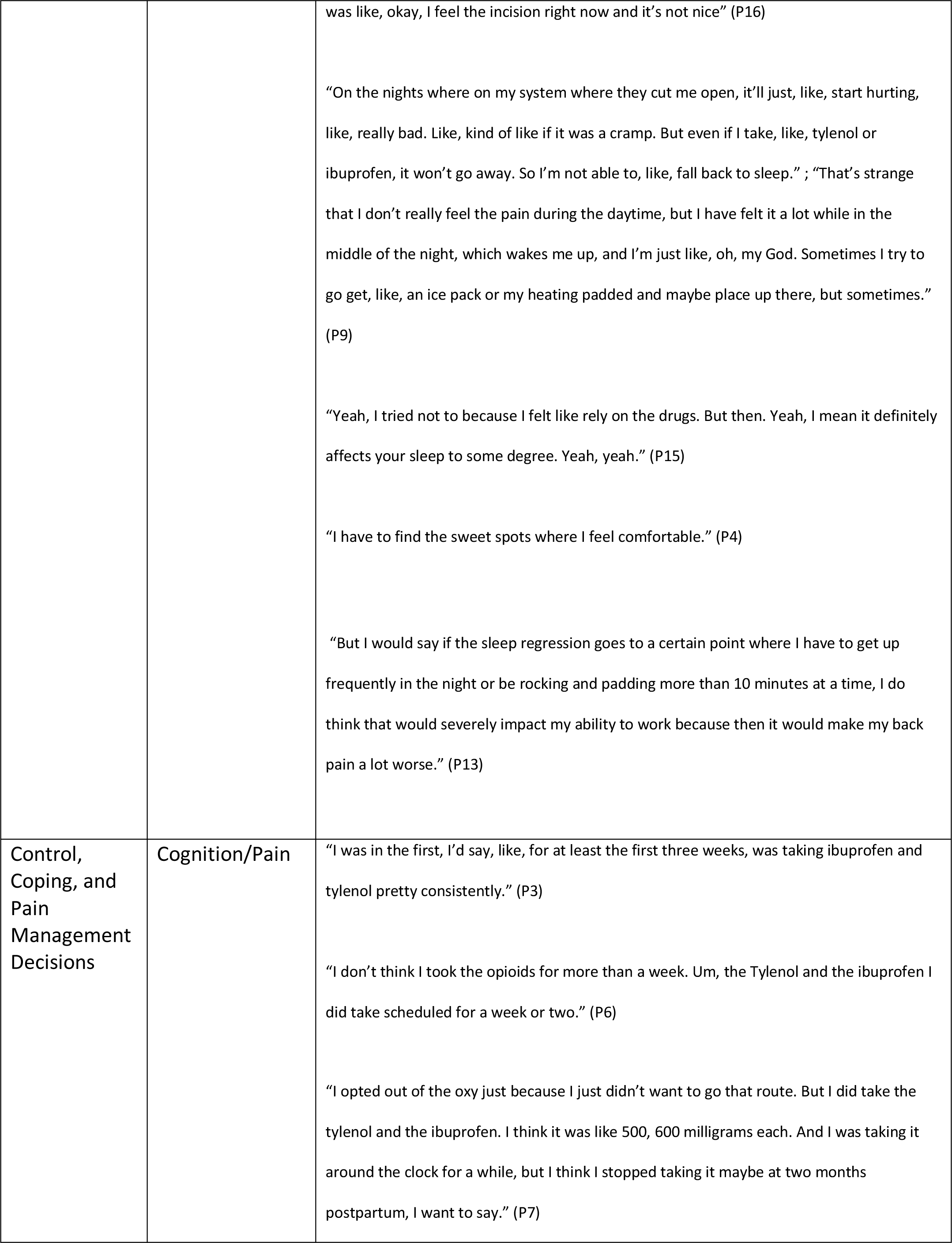
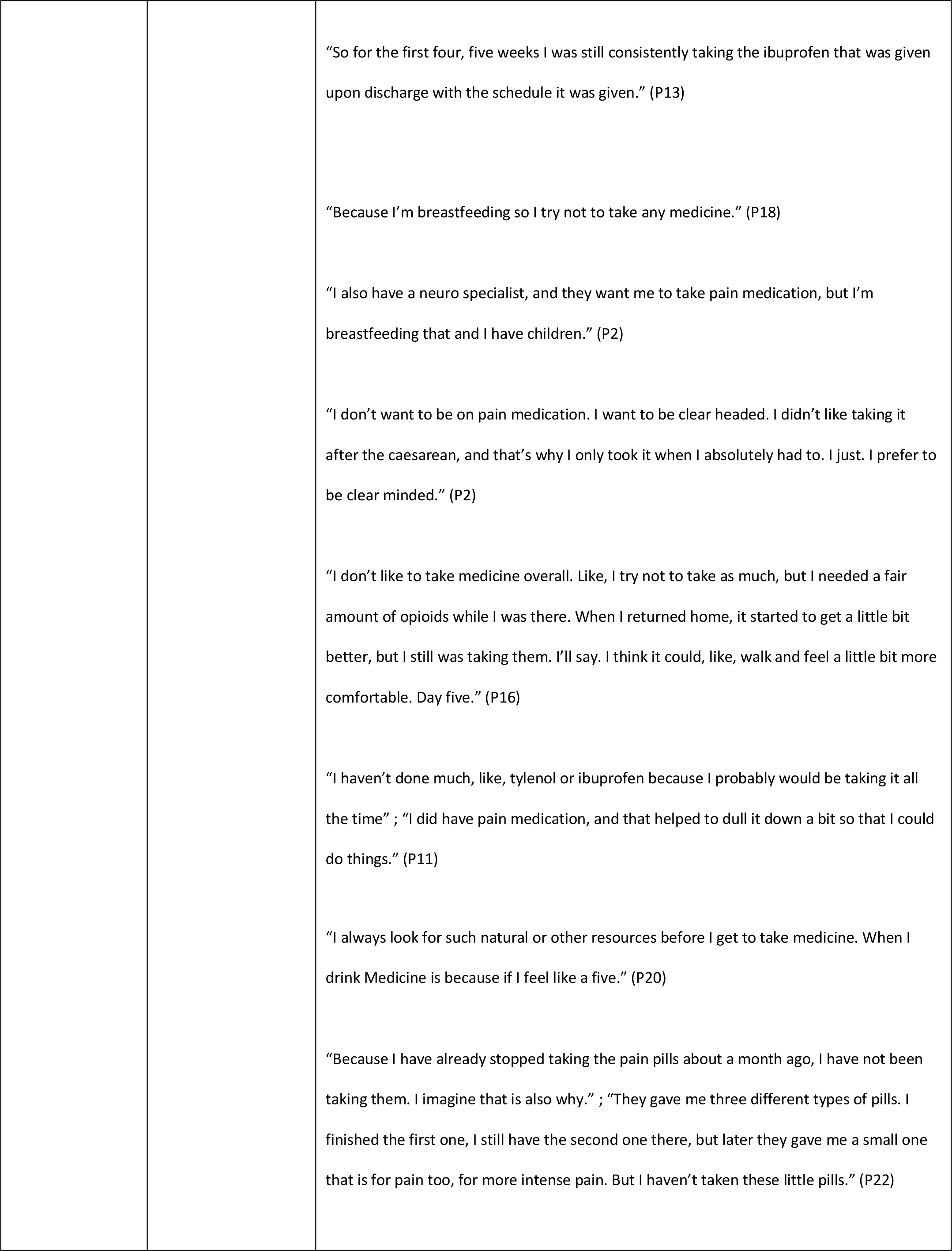
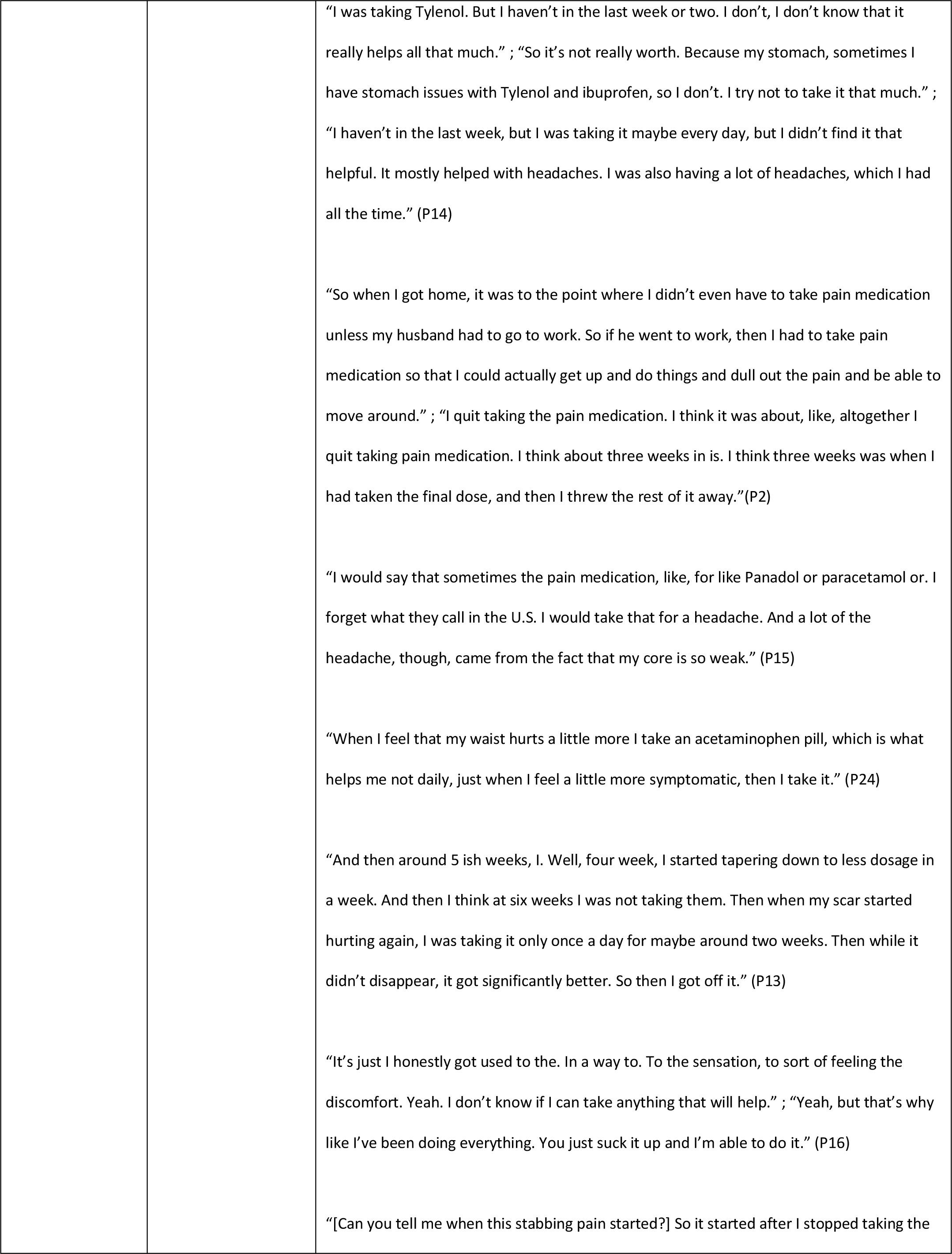
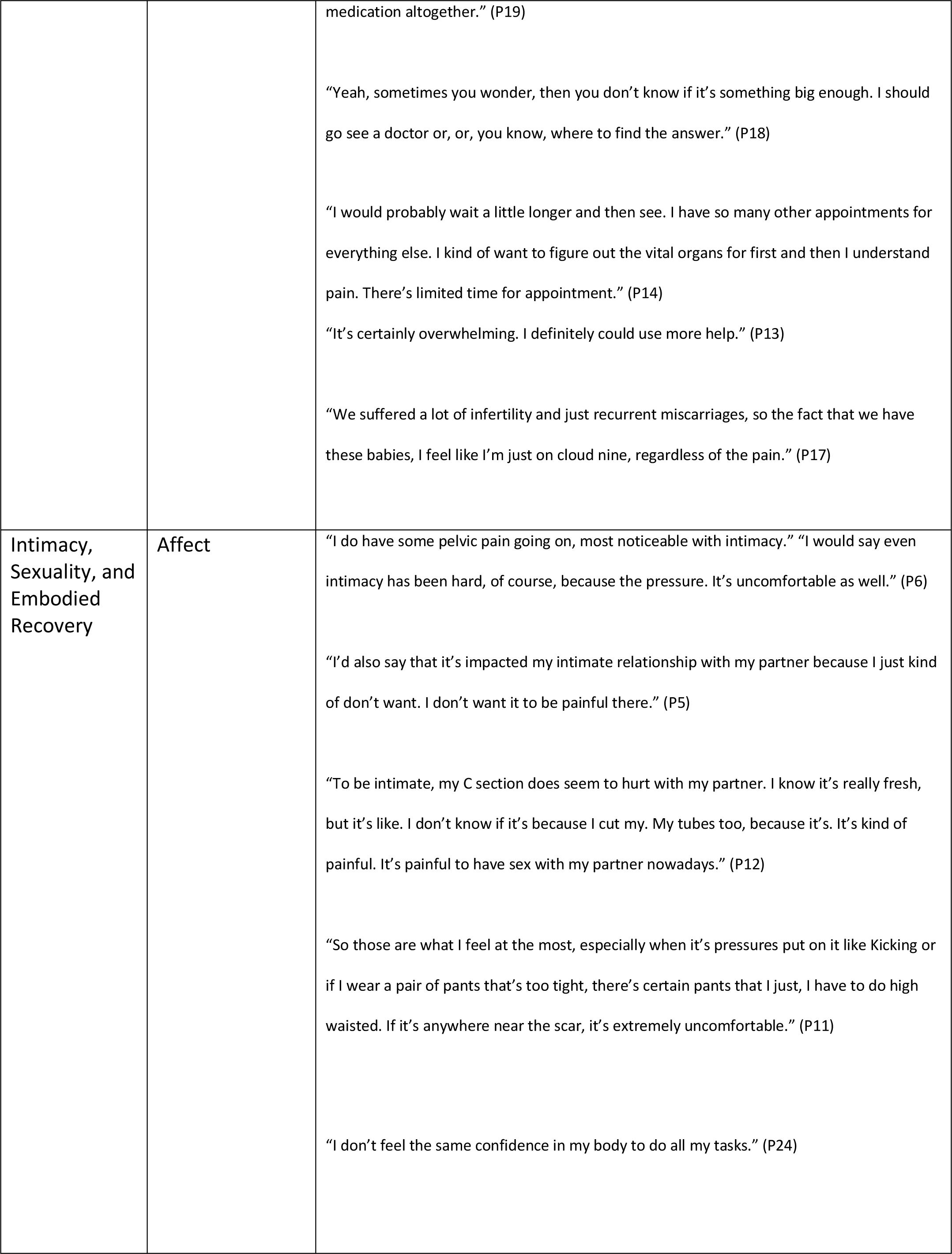
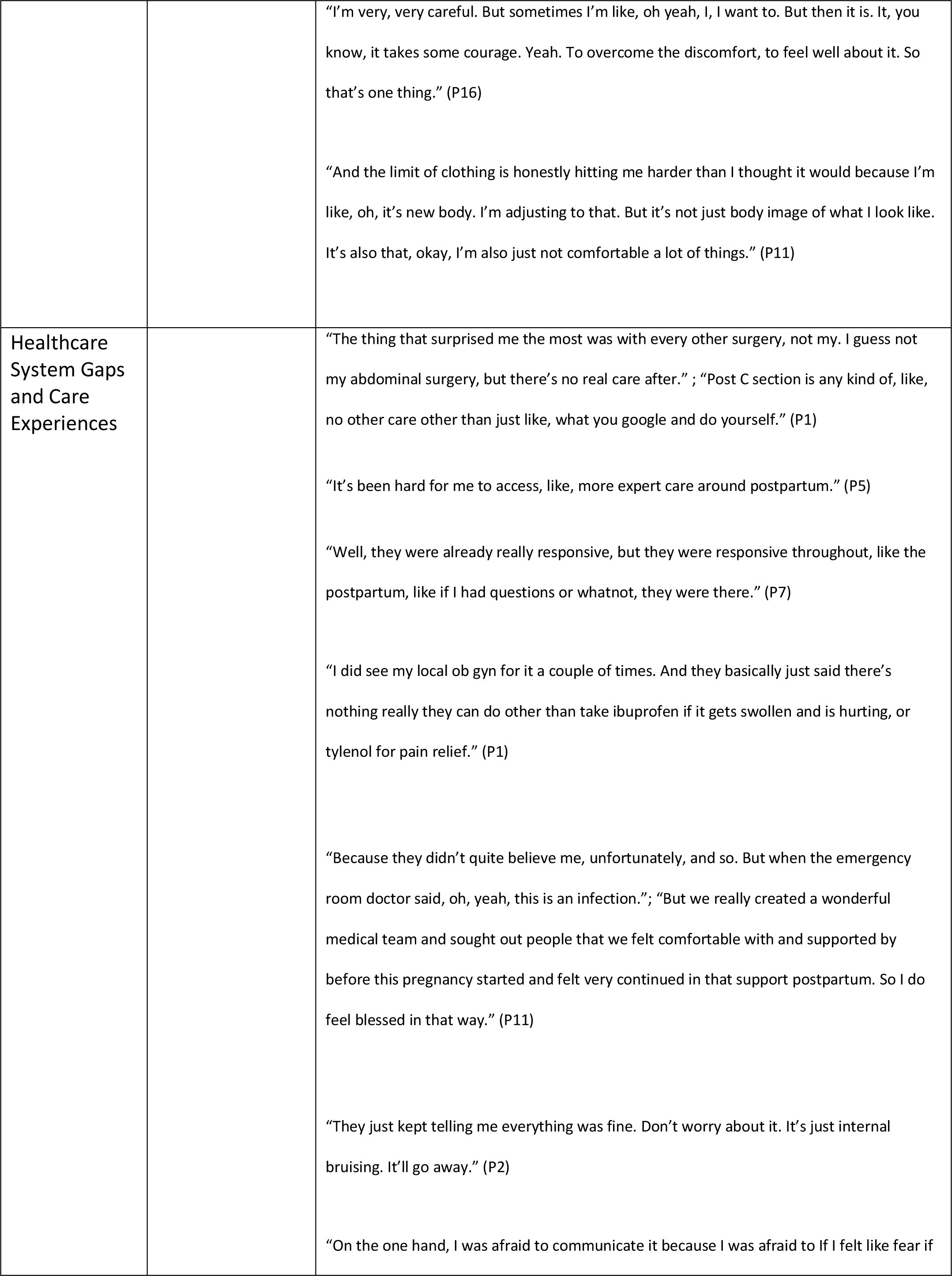
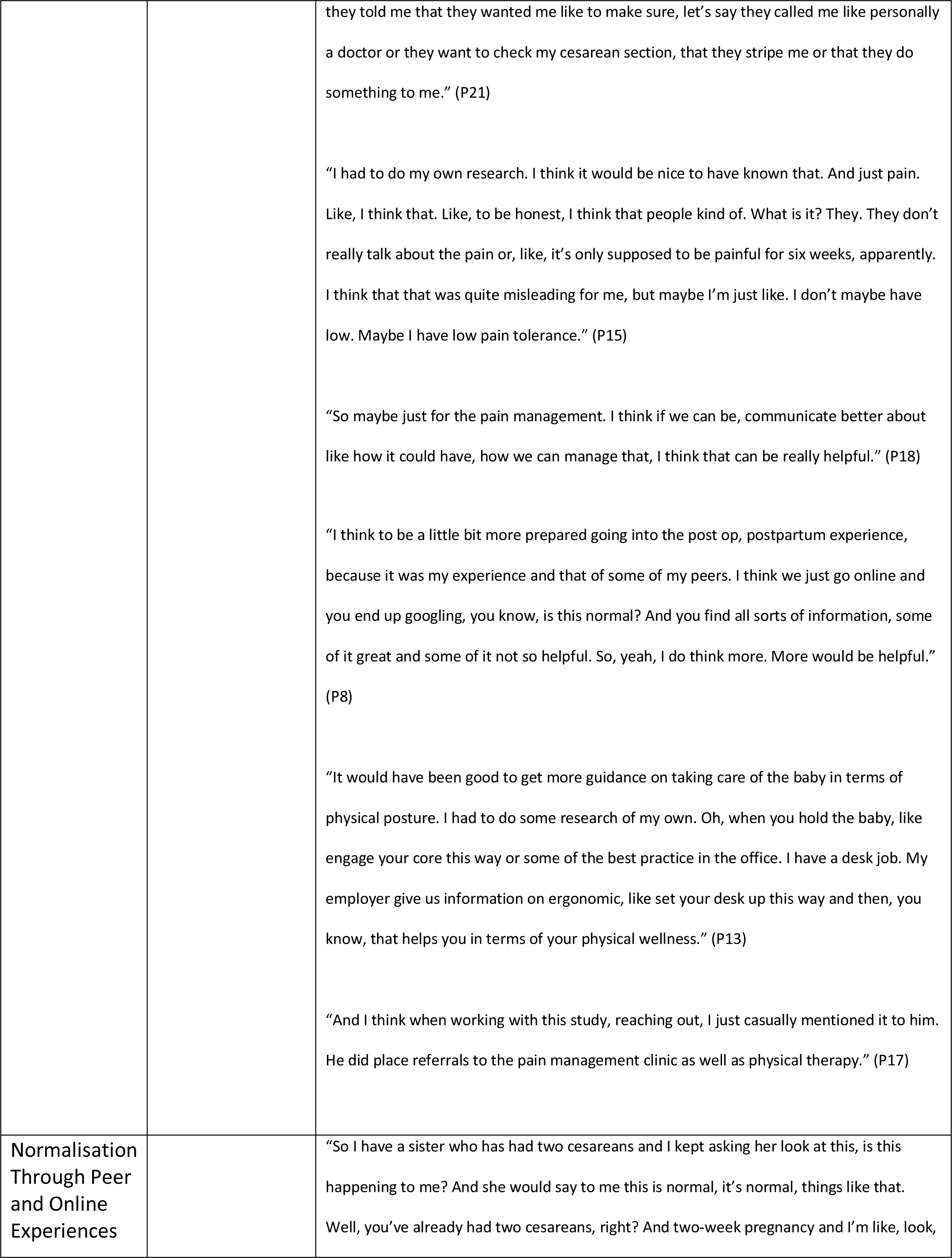
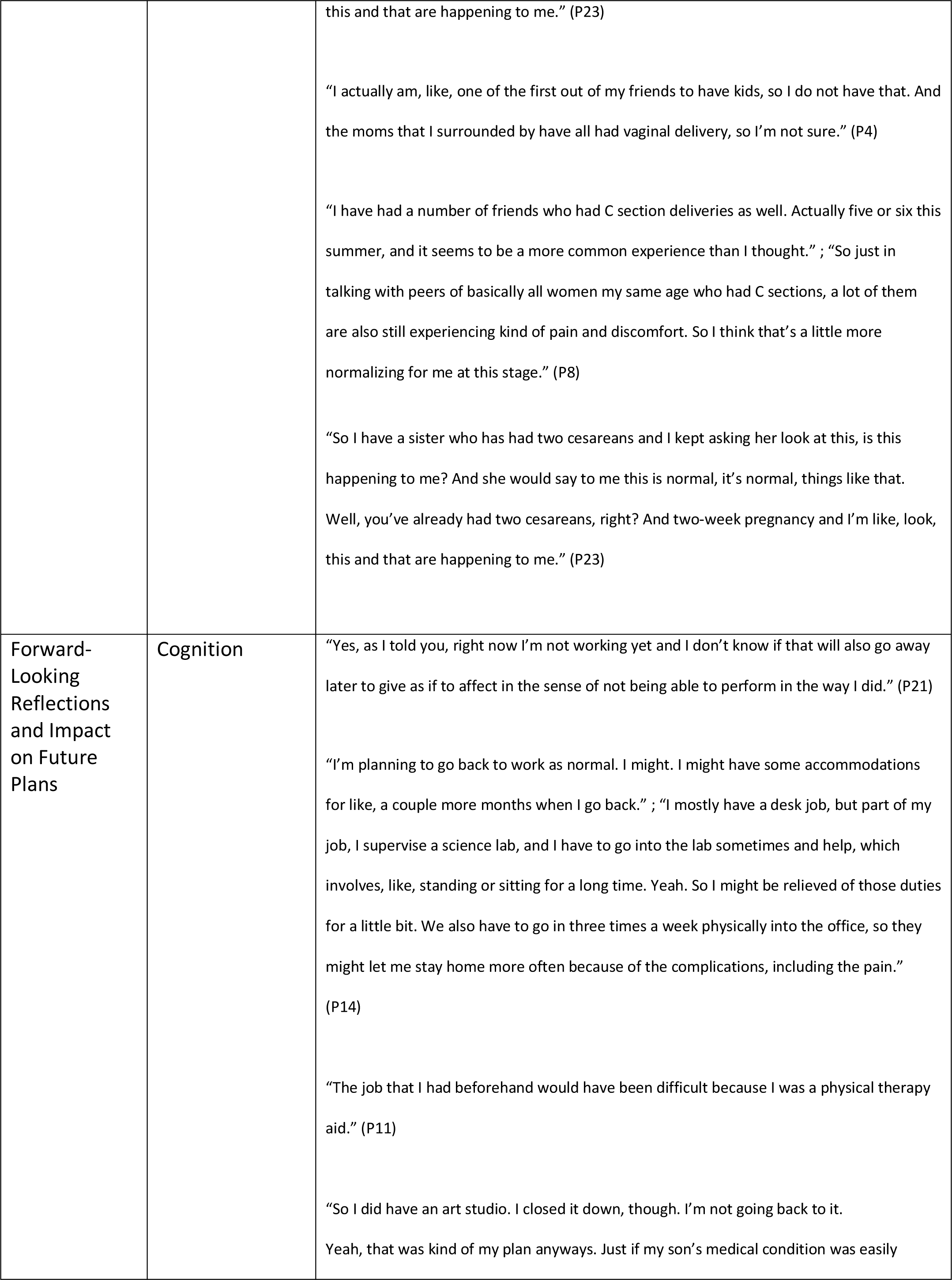
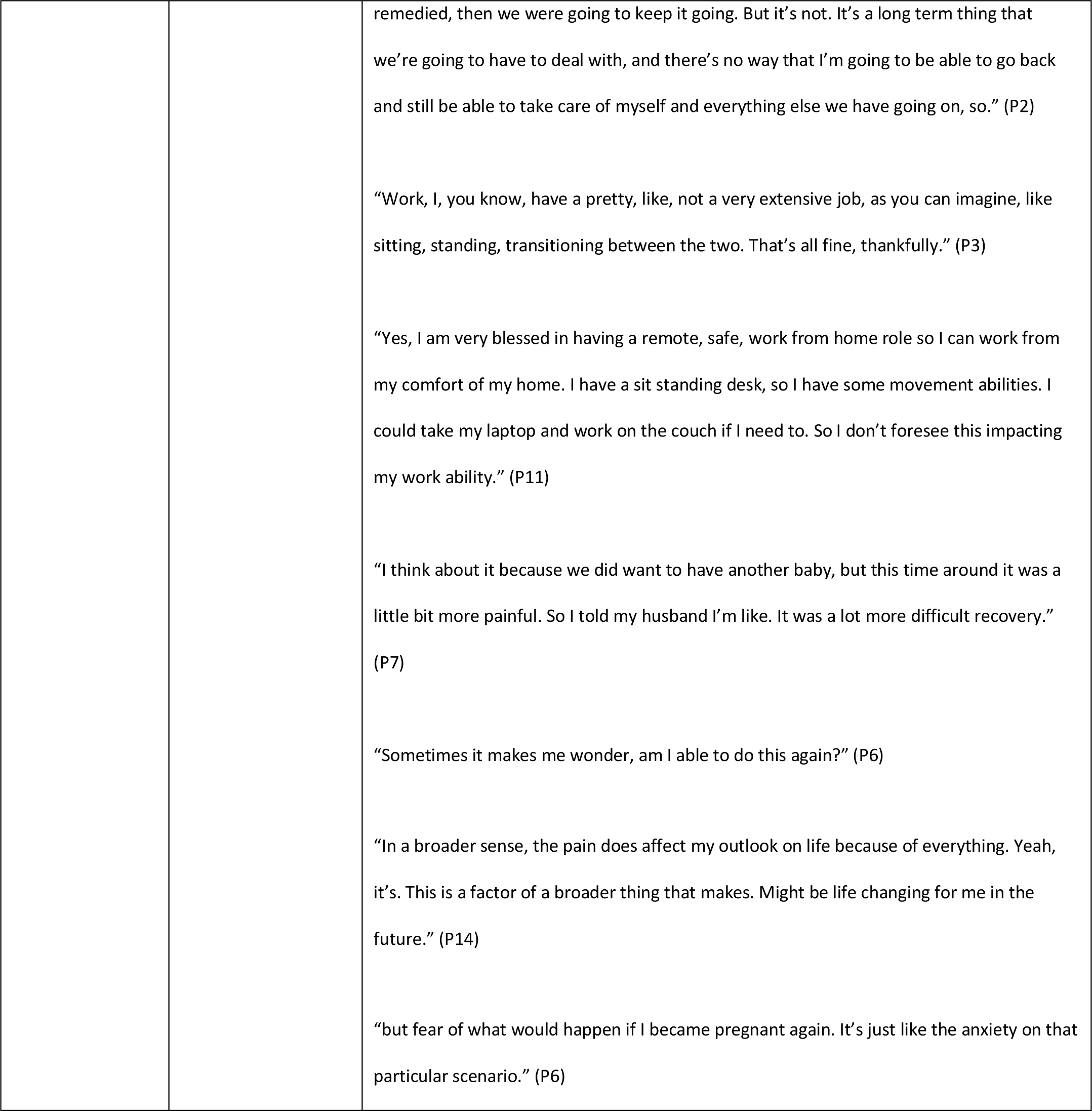
Exemplar quotes of postpartum pain symptoms, impacts, and coping strategies grouped according to themes and SPACE framework domains. Illustrative theme quotes. (PX) = Participant ID.

#### 2. Functional Limitations and Fatigue

Pain and reduced energy levels impacted participants’ ability to engage in physical activity and daily routines. Many adjusted their lifestyles to accommodate pain or avoid worsening. (Table 2).

Tasks such as lifting, walking, or standing for extended periods became challenging, and participants described becoming fatigued much more quickly than before delivery.

#### 3. Pain Interference with Infant Care and Identity

Pain hindered maternal caregiving and disrupted their transition to motherhood. Participants described physical limitations that restricted their ability to lift, feed, or soothe their infants. This interference often elicited distress, guilt, or a sense of inadequacy regarding their maternal role.

#### 4. Psychological Distress and Cognitive Load

Many participants described emotional distress, including anxiety, low mood, and self-doubt. Pain created uncertainty about healing and provoked fear that something was wrong.

These emotional and cognitive symptoms often compounded each other, especially in the absence of adequate support or reassurance.

#### 5. Sleep Disruption and Night-Time Vulnerability

Sleep was consistently affected by both pain and emotional arousal. Participants described difficulties falling asleep, staying asleep, and feeling rested. Sleep deprivation made daytime pain worse.

Pain at night was described as particularly distressing, contributing to emotional vulnerability and daytime fatigue.

#### 6. Control, Coping, and Pain Management Decisions

Pain management strategies varied widely, but many participants expressed reluctance to use strong analgesics, especially opioids. Concerns centred around potential side effects, the impact on maternal alertness, and transmission through breast milk.

Participants often opted for non-pharmacological methods, emphasising a desire for control and safety in the postpartum period.

#### 7. Intimacy, Sexuality, and Embodied Recovery

Chronic pain and body image concerns influenced participants’ relationships with their partners and their own bodies. Physical intimacy was often avoided due to pain or discomfort, and several women described feeling disconnected from their sense of self.

These embodied disruptions extended beyond physical symptoms, shaping self-esteem and relational dynamics.

#### 8. Healthcare System Gaps and Care Experiences

Participants commonly reported unmet needs in postpartum care. Many described inadequate follow-up, poor communication, and limited acknowledgement of their pain experiences.

There were some reported positive experiences, but more frequent reports of limited postpartum pain support by the healthcare system.

#### 9. Normalisation Through Peer and Online Experiences

Peer and online stories helped some participants validate their experiences, while others felt disheartened by comparisons. Shared narratives were a double-edged sword, providing reassurance or reinforcing isolation.

These interactions shaped perceptions of what constituted a ‘normal’ recovery and influenced help-seeking behaviour.

#### 10. Forward-Looking Reflections and Impact on Future Plans

Chronic pain prompted many to re-evaluate future plans, including decisions about childbearing, career, and personal resilience. For some, pain limited their aspirations; for others, it became a source of strength.

Participants reflected on how their recovery shaped their identity, values, and longer-term priorities.

Pain was often more intense and longer-lasting than expected. Two (8%) of women said they thought they were healing normally until pain worsened unpredictably weeks, or even months later. Lifting, bending, or holding their baby sometimes made things worse. This loss of physical function often came alongside low energy, and many felt stuck in a cycle: after doing too much, this triggered pain, and then the patient suffered exhaustion the following day. Patients described feeling frustrated, even ashamed, that they could not manage simple tasks. For ten patients (42%), this “weakness” impacted their recovery.

The emotional impact of CPSP was demonstrated by pain interference with maternal-infant bonding, which left two (8%) patients feeling guilty or inadequate, and in a few cases triggering anxiety or depressive symptoms. Five (21%) patients worried their body had not fully healed or feared tearing something internally. This constant hypervigilance added to the mental load of individuals. Sleep, already fragile in the postpartum period, was often made worse by pain. 13 (54%) patients said they lay awake, or that pain impacted them at night, leaving them more exhausted the next day. Over time, decreased total sleep time, pain, and low mood were related and negatively impacted each other. Two factors unique to the postpartum period, sleep disturbance and fatigue, are emerging as important drivers of persistent pain.

16 (67%) women had stopped or reduced analgesia dosing despite still experiencing pain. There was a real concern about taking too many analgesics and for too long, it was seen as a negative, either not the normal for postpartum recovery or it was not believed to be helpful for pain at that stage. These beliefs persisted even when scar pain returned. There was wide variation in dosing regimens used, with no guidance as to how to taper. 4 (16%) women made a conscious decision to avoid stronger pain relief, particularly opioids, because they were breastfeeding or wanted to stay clear-headed.

The sense of being “not myself anymore” came up in 4 (16%) patients. Not being believed or taken seriously by healthcare professionals, family, or peers, worsened this feeling.

8 (33%) women said that pain affected decisions about future pregnancies or work. A few left jobs or reduced their working hours or altered their mode of work. But not everyone framed their experience negatively. A small number described finding strength in having coped, or said that knowing their baby was healthy made it feel worthwhile.

## Discussion

Narratives from patient interviews revealed a multidimensional burden of chronic postsurgical pain, with persistent pain, sleep disruption, fatigue, psychological distress, and cognitive load interacting to impair infant care, functioning, and maternal identity. Ten themes captured physical, emotional, social, and healthcare-related impacts, with symptoms described as interrelated rather than isolated.

Women emphasised that pain was more than a physical sensation: it shaped maternal identity, interfered with caregiving, strained relationships, and influenced confidence and future plans. Sleep disturbance, fatigue, emotional distress, and cognitive load rarely occurred in isolation but interacted to worsen difficulties. Those who coped more effectively often reframed their experiences or developed strategies to break cycles of fear, pain, and exhaustion.

Sleep disturbance emerged as particularly important. One study of women with pregnancy-related back pain found that those with significant sleep disturbance at 4 months postpartum had nearly three time greater odds of experiencing persistent lumbopelvic pain compared to those sleeping well^9^. Pain and poor sleep likely reinforce each other in a cyclical manner, fragmenting rest and slowing healing.^10^ Postpartum fatigue, a prolonged state of exhaustion affecting many new mothers, further compounds this issue. Fatigue often correlates with emotional distress (including depressive symptoms) and both can increase pain sensitivity^11^. For many women, the combined effect of sleep loss and fatigue in caring for an infant can reduce normal pain tolerance.^12^

Reluctance to use analgesia was another recurring theme. Consistent with prior work^13^, many women tolerated moderate pain rather than risk perceived harms to the infant, whether from maternal drowsiness or medication transfer in breast milk. This underscores the need for clear, evidence-based guidance on safe postpartum analgesia and for interventions that address fears and misconceptions. Others talked about the toll of pain on relationships and intimacy. Some avoided sexual intercourse for fear of pain or felt disconnected from their bodies. A recent qualitative study of early recovery after elective CD found that women experienced significant pain in the first few days, followed by faster-than-expected improvement^14^. Consistent with our study, women expressed a clear need for better information, especially around what to expect physically and how to manage recovery at home.

Our interviews showed how symptoms, pain, fatigue, poor sleep, low mood, anxious thinking, rarely occurred in isolation. Our participants described a cycle that was hard to break. Those who coped better often had ways of reframing the experience or finding distraction, and did not describe fear, pain and exhaustion. Emerging evidence portrays the transition from acute to chronic postpartum pain as a biopsychosocial process.^15^ Beyond the physical surgical injury, a host of non-physical factors - social support, psychological health, and cognitive pain appraisals - critically shape recovery.^16^ Women lacking strong support networks (e.g. unpartnered mothers) report higher postpartum pain levels^17^, suggesting that social stressors can amplify pain perception. Psychological factors are particularly important. Pre-existing anxiety or depression has been associated with more intense and prolonged postpartum pain^18^. In a Chinese cohort, women with symptoms of depression before delivery were significantly more likely to develop CPSP up to 6 months after CD^18^. Similarly, a prior history of chronic pain (such as back pain) or higher acute pain on the first day postpartum increases the risk of long-term pain^11^. Pain catastrophizing, the tendency to have ruminating, negative thoughts about pain - is another key risk factor.^19^ This suggests that emotional state and pain-related cognitions can modulate pain experience and recovery course after childbirth.^20^

The study findings suggest we need to reconsider how pain recovery is approached after CD. Pain scores alone are increasingly considered insufficient to evaluate the risk of CPSP, and multidomain assessment is called for^21^. Domains such as sleep quality, mood, cognitive load and fatigue are highly relevant, and could be integrated into routine postpartum assessment.^22^ The Stanford Obstetric Recovery Checklist (STORK) has already demonstrated the value of broader, patient-centred recovery assessments in routine maternity care.^23^

Research indicates that poor sleep, intense acute pain, emotional distress and fatigue often cluster before chronic pain develops^24–27^. Pain severity correlates with poor sleep quality and emotional distress, mediated by anxiety about sleep and poor sleep hygiene^28^. Clusters of depressive symptoms, stress, fatigue, and lack of sleep in the perinatal period have been identified, which are potential contributors to adverse maternal health outcomes^29^. Another study also found the bidirectional association between sleep disturbance and pain, noting that sleep disturbance can lower pain thresholds and exacerbate psychological distress^30^. The importance of addressing sleep disturbance in chronic pain management has been highlighted, as poor sleep quality and insomnia-type symptoms are linked to higher levels of disability and depression^31,32^. Collectively, these studies suggest that addressing sleep disturbance and emotional distress early in the postpartum period could be crucial in preventing the development of chronic pain.

The SPACE-Postpartum framework (Sleep, Pain, Affect, Cognition, Energy; “SPACE”), was recently conceptualised through triangulation of qualitative interviews, cohort observations and published literature,^24,33–35^ with aim of comprehensively describing multidimensional postpartum pain and recovery. Key symptoms, impacts, and coping strategies identified in the current thematic analysis were consistent with the five pre-specified SPACE domains, thereby providing experiential support for this framework (Figure 1). Sleep disruption and fatigue were among the most frequently discussed concerns, described by almost all participants as worsening pain and distress, whereas cognitive impacts (concentration, intrusive thoughts) were less frequently discussed but strongly connected to mood disturbance. Although symptoms from all domains were described, pain interference, sleep disruption, and affective distress carried particular weight in narratives of daily functioning and maternal identity. Participants rarely described symptoms in isolation; rather, poor sleep worsened fatigue and mood, which in turn worsened pain, suggesting that relationships between domains were as important as the individual domains. In the present study, SPACE was applied only after inductive theme generation, serving as a means to integrate qualitative and quantitative findings, and to provide experiential validation of the framework. The convergence between participant-derived themes and the SPACE domains supports the model’s relevance, warranting evaluation in diverse populations and study designs.

We acknowledge that this study has limitations. This was a single-centre US study involving only 24 women, so findings may not reflect wider populations. While we included both English and Spanish speaking patients, other cultural and linguistic groups were not represented. As with all qualitative work, these are self-reported experiences that reflect individual perspectives. However data analysis was performed using robust methodology including independent coding and reflective analysis. SPACE was applied after inductive coding and theme development, in an interpretive manner. This carries some risk of deductive bias; however, all initial coding was inductive, and the framework was considered only once major thematic domains had been established. Its use was intended to support, rather than constrain, interpretation, consistent with a reflexive thematic approach.

In conclusion, this study demonstrates that CPSP after CD is rarely an isolated physical problem. Symptoms in sleep, mood, cognitive load, energy, and pain interact and impact each other, influencing both day-to-day function and 3-6 month pain experience. The themes identified aligned align with the multidomain SPACE-Postpartum framework, and support its further evaluation as a model for understanding and predicting postpartum pain outcomes.

## Data Availability

All data produced in the present study are available upon reasonable request to the authors

## Acknowledgements

We thank the participants who generously shared their experiences for this study.

## Author Contributions

**S.C.** conceptualised the SPACE-Postpartum framework, designed the study, conducted and analysed interviews, and drafted and revised the manuscript.

**P.C.** contributed to qualitative analysis and critical manuscript revision.

**G.M.** conducted Spanish interviews and supported qualitative analysis and manuscript revision.

**D.P.** assisted with data collection and manuscript editing.

**B.C.** provided project supervision, methodological feedback and reviewed the manuscript.

**P.S.** provided project supervision, methodological feedback, qualitative analysis input, and reviewed the manuscript.

## Conflicts of Interest

This study received partial support from a Stanford University research grant (Prof. Sultan). S.C. reports a UK provisional patent application related to the SPACE framework. Other authors declare no competing interests.

## Declaration of Generative AI and AI-assisted technologies in the writing process

During the preparation of this work the author(s) used ChatGPT to assist with refining wording and improving clarity. After using this tool/service, the author(s) reviewed and edited the content as needed and take(s) full responsibility for the content of the publication. All coding, thematic analysis, and interpretation of qualitative data were conducted independently by the authors.

## Notes

### Competing Interest Statement

S.C is named on a UK provisional patent application related to the SPACE framework

### Author Declarations

Stanford University School of Medicine IRB approved

